# Visit-to-visit blood pressure variability and progression of white matter hyperintensities over 14 years

**DOI:** 10.1101/2023.03.02.23286727

**Authors:** Esther Janssen, Jan-Willem van Dalen, Mengfei Cai, Mina A. Jacob, José Marques, Marco Duering, Edo Richard, Anil M. Tuladhar, Frank-Erik de Leeuw, Nina Hilkens

## Abstract

There is evidence that blood pressure variability (BPV) is associated with cerebral small vessel disease (SVD) and may therefore increase the risk of stroke and dementia. It remains unclear if BPV is associated with SVD progression over years. We examined whether visit-to-visit BPV is associated with White Matter Hyperintensity (WMH) progression over 14 years and MRI markers after 14 years.

We included participants with SVD from the Radboud University Nijmegen Diffusion tensor Magnetic resonance imaging Cohort (RUNDMC) who underwent baseline assessment in 2006 and follow-up in 2011, 2015 and 2020. BPV was calculated as coefficient of variation of BP at all visits. Association between WMH progression rates over 14 years and BPV was examined using linear-mixed effects model. Regression models were used to examine association between BPV and MRI markers at final visit in participants.

A total of 199 participants (60.5 SD 6.6 years) who underwent four MRI scans and blood pressure measurements were included, with mean follow-up of 13.7 (SD 0.5) years. Systolic BPV was associated with higher progression of WMH (β = 0.013, 95% CI 0.005 – 0.022) and higher risk of incident lacunes (OR: 1.10, 95% CI 1.01-1.21). There was no association between systolic BPV and grey and white matter volumes, Peak Skeleton of Mean Diffusivity (PSMD) or microbleed count after 13.7 years.

Visit-to-visit systolic BPV is associated with increased progression of WMH volumes and higher risk of incident lacunes over 14 years in participants with SVD. Future studies are needed to examine causality of this association.

## Introduction

Damage to the small arteries and arterioles of the brain is commonly referred to as cerebral small vessel disease (SVD).^1^ SVD is the most important vascular contributor to dementia, causes the majority of intracerebral haemorrhage and about one quarter of all ischemic strokes, but its pathophysiology remains poorly understood.^1,2^ As pathology of these arterioles cannot be visualized in vivo with conventional 3T Magnetic Resonance Imaging (MRI), MRI markers of SVD including white matter hyperintensities (WMH), lacunes and microbleeds, as well as reductions in white matter microstructural integrity, are considered to reflect the underlying pathology. ^3^

Although hypertension is the most established risk factor for SVD, studies examining the effects of lowering blood pressure (BP) showed mixed results.^4^ The SPRINT-MIND trial has demonstrated that intensive BP reduction can reduce WMH progression, but this effect seems modest.^5^ Moreover, trials of BP lowering drugs do not show a consistent reduction in risk of incident dementia or stroke.^6,7^ Emerging evidence suggests that other BP measures, such as blood pressure variability (BPV) over a period of hours, days or years hold additional prognostic value.^8^ Higher BPV independent of mean BP was found to be associated with increased mortality,^9^ higher dementia risk^10^ and increased risk of stroke and other cardiovascular events.^11^

In patients with SVD, higher systolic BPV is associated with higher burden of WMH and progression of WMH. ^12,13^ Most of the previous studies only assess SVD burden at one time-point. However, it is crucial to determine if BPV is merely a consequence of SVD or a causal factor by studying incident SVD or progression of SVD. To date, studies that assessed progression of SVD markers only measure progression over a short time interval, which is unlikely to capture the long-term temporal dynamics of SVD progression.^14,15^ If BPV is associated with progression of MRI markers of SVD over a longer period of time beyond mean

BP levels, a next step would be to investigate if reducing BPV results in prevention or mitigation of SVD in patients at risk. This study aims to examine the effects of visit-to-visit BPV on progression of WMH volumes over 14 years and SVD burden after 14 years.

## Methods

### Study population

This study is part of the RUN DMC study, a single-center prospective cohort study of 503 individuals with SVD, described in detail previously.^16^ In short, this study aims to examine the risk factors, progression and clinical consequences of SVD.^16^ Patients aged 50-85 years who were referred to Radboud University Medical Center Neurology outpatient clinic between October 2002 and November 2006 were screened for inclusion. SVD diagnosis was based on MRI and included presence of WMH and/or lacunes. Exclusion criteria were presence of dementia and/or parkinsonism, other (psychiatric) diseases interfering with testing or follow-up, SVD mimics (i.e. multiple sclerosis), a life expectancy <6 months or contraindications for MRI. The Medical Review Ethics Committee region Arnhem-Nijmegen approved the study and all participants gave written informed consent.

Of the 503 participants at visit 1, 361 had an MRI at visit 2 (mean interval of 5.4 years), 296 at visit 3 (mean interval of 3.4 years) and 231 at visit 4 (mean interval of 5.0 years). Patients had more BP measurements available than MRI scans, due to MRI contraindications in the elderly population (Figure 1). Because missing BP measurements would lead to large intervals between BP measurements, participants were only included in this analysis if BP measurements and MRI data were available at all four visits (n=199). A flowchart of the study population over time is shown in Figure 1.

**Figure 1:**
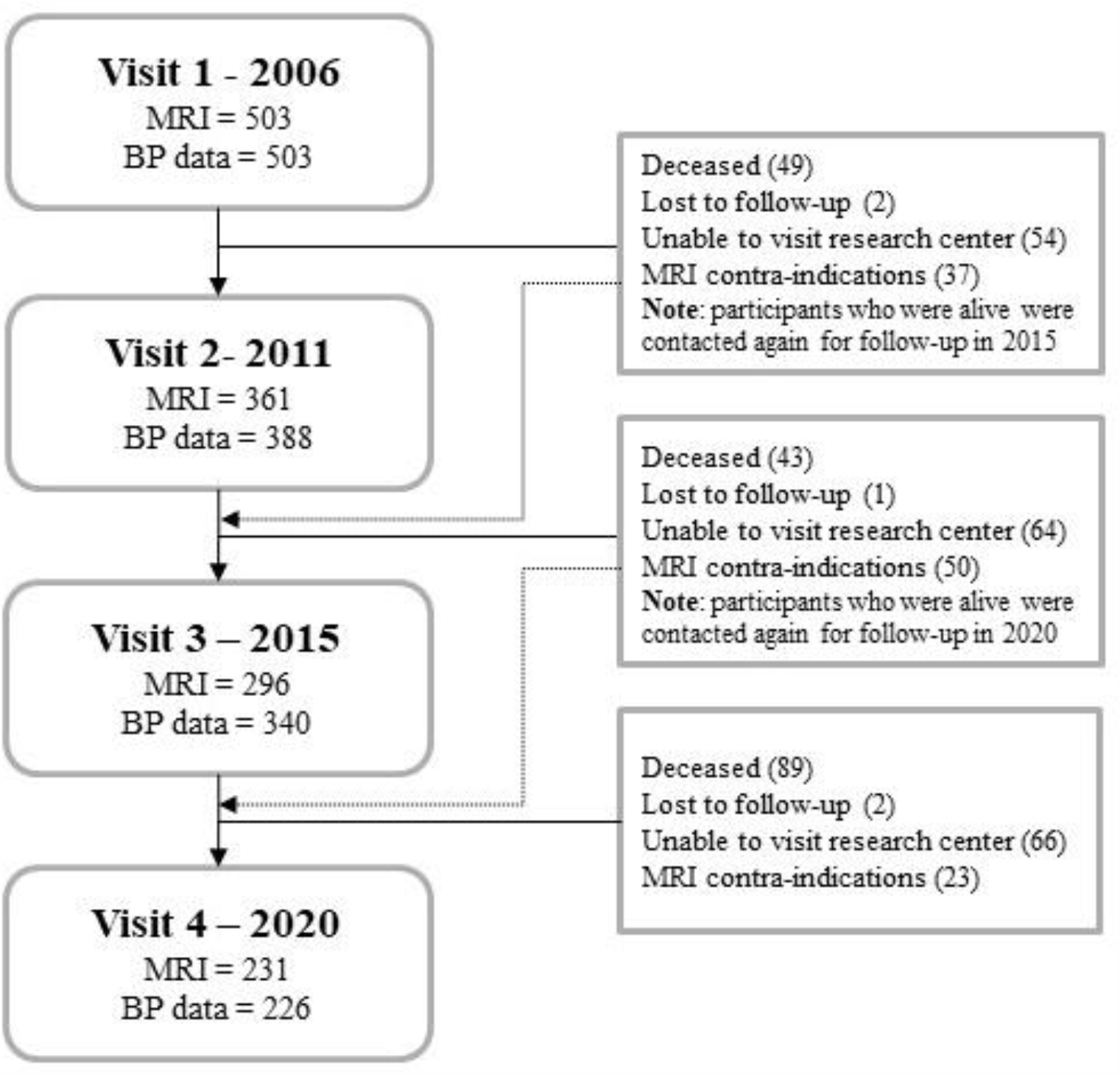
Flowchart of included participants at all study visits. Participants who were unable to visit research center and were alive between 2011-2015 and 2015-2020 were contacted again for next follow-up (as shown by dotted line).

### Blood pressure assessments and calculation of BP variability

At every study visit, BP was recorded as the mean of three measurements in supine position after five minutes of rest, using an automated oscillometric device (CARESCAPE V100, GE Dinamap, USA). BP was measured at four visits over 13.7 years in all participants included in this analysis. BPV was defined as the Coefficient of Variation (CV) of the systolic pressure over all visits, calculated as the standard deviation divided by the mean BP, multiplied by 100%. Both systolic and diastolic BPV were calculated separately. Information about current antihypertensive medication use was obtained at each visit.

### MRI protocol

Images were acquired on 1.5T MRI at first three visits (in 2006: Siemens, Magnetom Sonata, in 2011 and 2015: Siements, Magnetom Avanto) and 3T at visit 4 (Siemens, Magnetom Prisma). In short, brain scans included the following sequences: 3D T1 Magnetization Prepared Rapid Gradient Echo (MPRAGE), 3D fluid-attenuated inversion recovery (FLAIR) and a transversal T2*-weighted gradient echo. At visit 4, a Susceptibility-Weighted Image (SWI) was derived from magnitude and phase data from the multi-echo gradient sequence. The acquisition details have been published previously.^17^ During visit 4 in 2020, the protocol included diffusion weighted imaging (DWI) consisted of diffusion-encoding directions: 10 x b=0 s/mm^2^, 30 x b=1000 s/mm^2^ and 60 x b=3000 s/mm^2^.

### MRI processing and brain volumetry

At visit 1, 2 and 3, WMH volumes were calculated using a semi-automated detection method using FLAIR and T1 sequences that was described previously.^18^ At visit 4, MP2RAGE data were processed to obtain T1-weighted images using code freely available online (https://github.com/JosePMarques/MP2RAGE-related-scripts). Registered and bias-corrected T1 and FLAIR images were used to segment WMH using a 3D U-net deep learning algorithm.^19^ All WMH segmentations were checked for and cleaned from artifacts using a custom 3D editing tool in Matlab (R2018a, The Mathworks, Natick, MA). Moreover, follow-up FLAIR images were aligned to baseline FLAIR images using FMRIB’s Linear Image Registration Tool (FLIRT) so all FLAIR scans have the same voxel size and slice thickness. This was done to minimize the effects of changes in FLAIR acquisition parameters during follow-ups. WMH volumes during follow-ups were corrected to baseline Intracranial Volume (ICV) to correct for interscan differences and normalized to baseline ICV.

Brain volumes, including grey matter volume (GM), white matter volume (WM) and cerebrospinal fluid volume (CSF) were calculated using SPM12 on the T1 MPRAGE images that were corrected for WMH. ICV was calculated by summing up GM, WM and CSF volumes. We corrected for differences in ICV between baseline and follow-up to account for MRI scanner changes.

### MRI markers of SVD

SVD markers (i.e. WMH volume, lacune count and microbleed count) were rated using Standards for ReportIng Vascular changes on nEuroimaging (STRIVE) criteria.^3^ Two trained raters manually rated lacunes on T1 and FLAIR scans and microbleeds on SWI scans, followed by a consensus meeting. To improve identification of incident lacunes during follow-ups, difference images for T1 and FLAIR sequences were used.^20^ First, the Brain Extraction Tool (BET) in FSL was used to create skull stripped images and image intensity was normalized to the 95% percentile. After this, follow-up images were registered to the baseline MRI scans. These baseline images were then subtracted from the registered and intensity-normalized follow-up images to create difference images. Incident lacunes are visible as hypointense voxel clusters on a uniform background, making it easier to identify them. Lacune count during follow-ups was calculated as the sum of the baseline lacune count and the number of incident lacunes for each subject.

### Diffusion Tensor Imaging (DTI) analysis

To examine microstructural integrity, we used the DTI scans at visit 4 to calculate the Peak width of Skeletonized Mean Diffusivity (PSMD). This is a robust imaging marker to capture global SVD-related microstructural damage in main white matter tracts.^21^ This was done using a fully automated pipeline (available at https://github.com/miac-research/psmd). ^21^

### Statistical analysis

Differences in baseline characteristics between participants included and not included in this analysis were examined using Mann-Whitney test or chi-square test where appropriate. We used multivariable linear regression to assess association between BPV and continuous outcomes at final visit (GM/WM volume, PSMD, WMH volume, all log-transformed due to non-normal distribution). Models were adjusted for baseline age, sex, mean BP (either systolic in models assessing association with systolic BPV or diastolic in models examining association with diastolic BPV) and use of antihypertensive medication at baseline. In a second model assessing the association between BPV and WMH volume, we additionally adjusted for baseline WMH volume. Separate models were used to examine effects of systolic and diastolic BPV. Because the changes in scanner system likely had a profound effect on volumetric measures (i.e. GM/WM volumes) and DTI measures (i.e. PSMD), we were not able to adjust models assessing these outcomes for baseline burden. To examine if BPV is associated with microbleed count at visit 4, we used negative binomial regression. This can be used to model overdispersed count outcomes with excess zeroes, such as microbleed count. Model was adjusted for baseline age, sex, mean BP (either systolic in models assessing association with systolic BPV or diastolic in models examining association with diastolic BPV) and use of antihypertensive medication. Negative binomial regression was also used to examine the relation between BPV and incident lacune count, adjusting for age, gender, mean BP (systolic or diastolic, depending on model), use of antihypertensive medication and baseline lacune count.

Furthermore, linear-mixed effects (LME) models were used to examine WMH progression. Since LME models allow modelling of subject-specific response trajectories over time, this can be used to examine changes in continuous outcomes in the entire population while accounting for individual differences in trajectories. To calculate individual progression rates per participant, we used a base model with follow-up time and baseline age as fixed effects and follow-up time as the random effect for each participant (model A). We then extracted the random slopes of follow-up time for each participant to examine the individual WMH progression rates.

Separate models were used to assess the effects of systolic and diastolic BPV. In the LME models used to examine the effect of BPV on WMH progression, we included the following fixed effects: systolic or diastolic BPV, baseline age, sex, baseline WMH volume, mean BP (systolic or diastolic, depending on model), use of antihypertensive medication at baseline, MRI scanner system and an interaction term between time and BPV (model B). We examined the effects of BPV using both continuous and quartile-based categorical scales in separate models. We evaluated change in Akaike information criterion (AIC) by one-way ANOVA to compare model fit with and without random intercepts and slopes per subject. Moreover, we compared model fit between the full model and the full model with a quadratic term to evaluate non-linear progression of WMH volume.

One of the assumptions of LME models is (approximately) normal distribution of residuals. In models where this assumption was not met, non-parametric bootstrap method with 1000 samples was performed to calculate 95% confidence intervals (CIs) and to infer p-values. Statistical analyses were performed with R version 4.1.0.

## Results

### Baseline characteristics

Of the 503 participants in the RUNDMC study, 199 were included in this study (Figure 1). Mean follow-up time until visit 2 was 5.3 (SD 0.2) years, 8.7 (SD 0.2) years until visit 3 and 13.7 (SD 0.5) years until visit 4. Baseline characteristics of the included and excluded participants are shown in Table 1. Participants included in the present study were younger at baseline than nonparticipants (60.5 SD 6.6 vs 68.0 SD 8.5) and had less vascular risk factors (i.e. diabetes, hypercholesterolemia, hypertension). Moreover, included participants had lower WMH volumes and less lacunes at baseline. The mean systolic and diastolic BP levels at baseline were 133.9 (SD 17.5) mmHg and 78.1 (SD 9.1) mmHg. In the included participants, mean systolic BPV was 9.9 (SD 4.9) and mean diastolic BPV was 9.6 (SD 3.9).

**Table 1.**
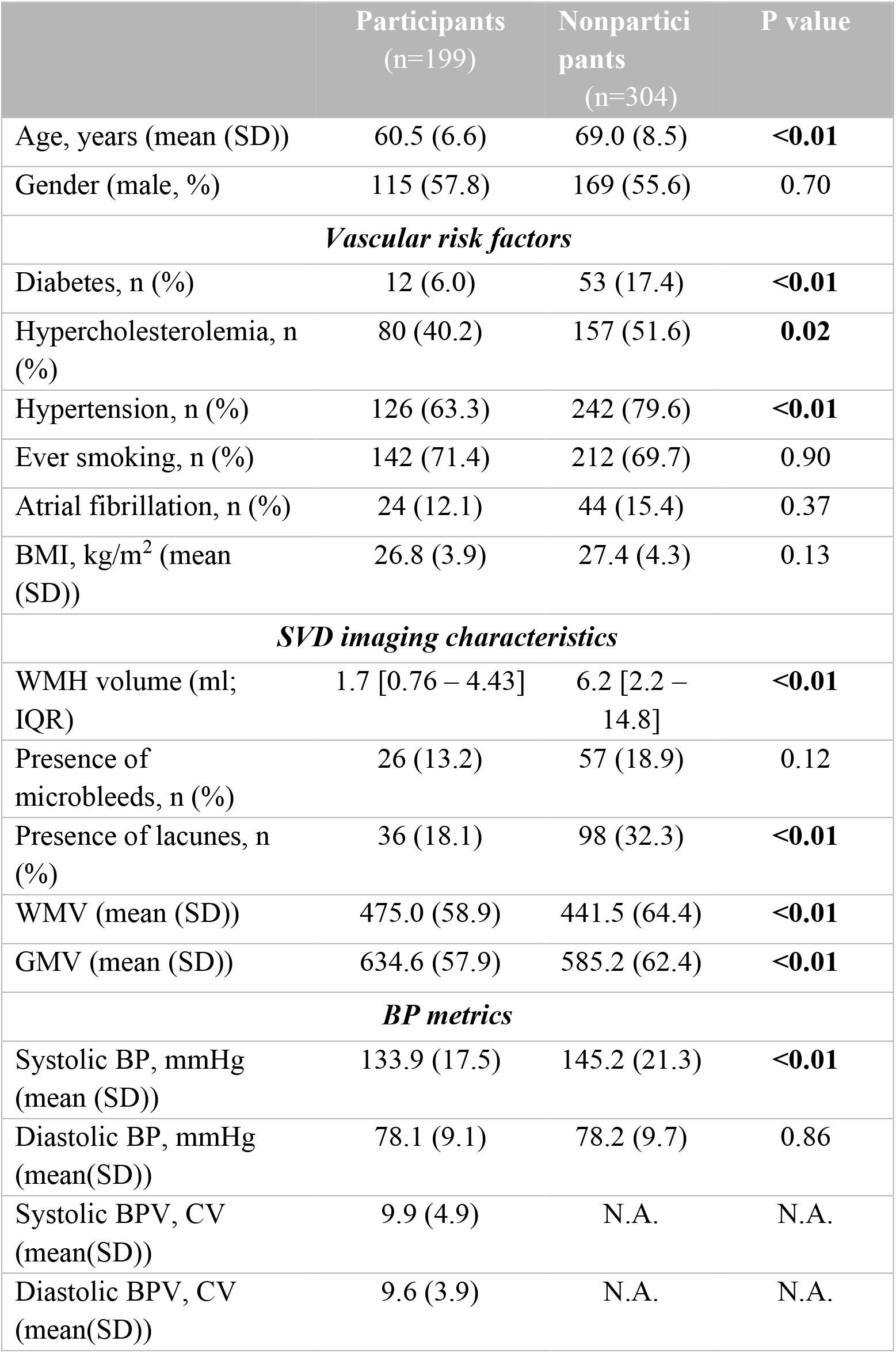
Baseline characteristics of participants included and not included in analysis. Data represent means (SD) and numbers (%). BMI: Body Mass Index, BP: Blood Pressure, BPV: Blood Pressure Variability, CV: Coefficient of Variation, GMV: Grey Matter Volume, WMV: White Matter Volume, WMH: White Matter Hyperintensity.

|  | Participants<br>(n=199) | Nonpartici<br>pants<br>(n=304) | P value |
| --- | --- | --- | --- |
| Age, years (mean (SD)) | 60.5 (6.6) | 69.0 (8.5) | **<0.01** |
| Gender (male, %) | 115 (57.8) | 169 (55.6) | 0.70 |
| ***Vascular risk factors*** |  |  |  |
| Diabetes, n (%) | 12 (6.0) | 53 (17.4) | **<0.01** |
| Hypercholesterolemia, n (%) | 80 (40.2) | 157 (51.6) | **0.02** |
| Hypertension, n (%) | 126 (63.3) | 242 (79.6) | **<0.01** |
| Ever smoking, n (%) | 142 (71.4) | 212 (69.7) | 0.90 |
| Atrial fibrillation, n (%) | 24 (12.1) | 44 (15.4) | 0.37 |
| BMI, kg/m<sup>2</sup> (mean (SD)) | 26.8 (3.9) | 27.4 (4.3) | 0.13 |
| ***SVD imaging characteristics*** |  |  |  |
| WMH volume (ml; IQR) | 1.7 [0.76 – 4.43] | 6.2 [2.2 – 14.8] | **<0.01** |
| Presence of microbleeds, n (%) | 26 (13.2) | 57 (18.9) | 0.12 |
| Presence of lacunes, n (%) | 36 (18.1) | 98 (32.3) | **<0.01** |
| WMV (mean (SD)) | 475.0 (58.9) | 441.5 (64.4) | **<0.01** |
| GMV (mean (SD)) | 634.6 (57.9) | 585.2 (62.4) | **<0.01** |
| ***BP metrics*** |  |  |  |
| Systolic BP, mmHg (mean (SD)) | 133.9 (17.5) | 145.2 (21.3) | **<0.01** |
| Diastolic BP, mmHg (mean(SD)) | 78.1 (9.1) | 78.2 (9.7) | 0.86 |
| Systolic BPV, CV (mean(SD)) | 9.9 (4.9) | N.A. | N.A. |
| Diastolic BPV, CV (mean(SD)) | 9.6 (3.9) | N.A. | N.A. |

### Association between BPV and WMH

Median WMH volume of all participants was 1.7 ml (Interquartile Range(IQR) 0.8 – 4.4) at baseline and 4.5 ml (IQR 1.7 – 11.3) at visit 4. Systolic BPV over all visits is associated with higher WMH volumes at baseline (standardized β = 0.18, p = 0.01), especially in the participants in the highest systolic BPV quartile (standardized β = 0.27, p = 0.001). When dividing the participants in quartiles based on systolic BPV, median WMH volume at visit 4 was 2.5, 4.5, 4.6, 7.2 ml for systolic BPV Q1, Q2, Q3 and Q4. BPV was only associated with WMH volume at 13.7 years in the participants in the highest systolic BPV quartile (standardized β = 0.21, p = 0.01) but not in the whole study population (standardized β = 0.12, p = 0.07) when adjusting for sex, age, mean systolic BP and use of antihypertensive medication. After additional adjustment for baseline WMH volume, this association was no longer significant in Q4 (standardized β = 0.12, p = 0.07).

The median annualized progression rate per quartile, estimated using LME, was 0.11, 0.21, 0.18, 0.29 mL/year for systolic BPV Q1, Q2, Q3 and Q4 (Model A; Figure 2). Both random intercept and random slope led to the best model fit for the mixed-effects model, while including a quadratic term did not significantly improve model fit. Therefore, the model without quadratic terms was used as the final model. The mixed-effects model of WMH change over time showed an overall interaction between follow-up duration and systolic BPV on WMH volume (β = 0.013, CI: 0.006 – 0.022), meaning that one point increase in CV of systolic BP was significantly associated with 0.013 ml/y [95% CI 0.006 – 0.022] WMH progression (Model B). The rate of WMH change was significantly higher in participants with high systolic BPV (Q4) compared to participants with low systolic BPV (Q1) (β = 0.239, CI: 0.126 – 0.410) (Figure 2/Table 2), meaning that in participants with high BPV, WMH volume increases with 0.239 ml/y more than in participants with low BPV. Diastolic BPV was not associated with WMH progression (Table S1/Figure S1).

**Table 2.**
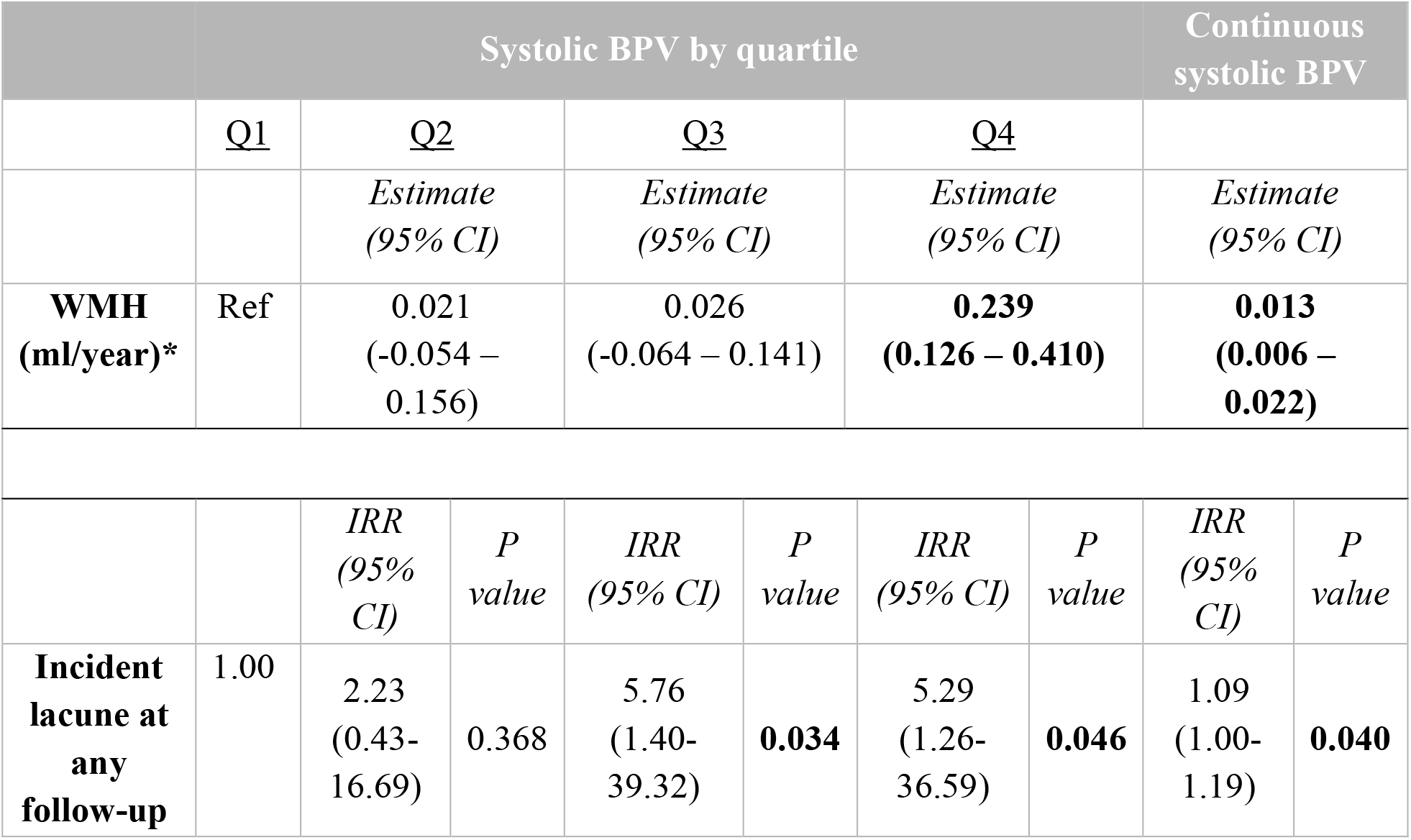
Association between systolic BPV and progression of WMH and incident lacunes. Data represent β estimates (95% CI), IRR values (95% CI) or p-values. Model was adjusted for sex, age, baseline WMH volume, use of antihypertensive medication at baseline, mean systolic BP and MRI scanner system. *Bootstrapped 95% CIs. BPV: Blood Pressure Variability, IRR: Incidence Rate Ratio, WMH: White Matter Hyperintensity.

**Figure 2.**
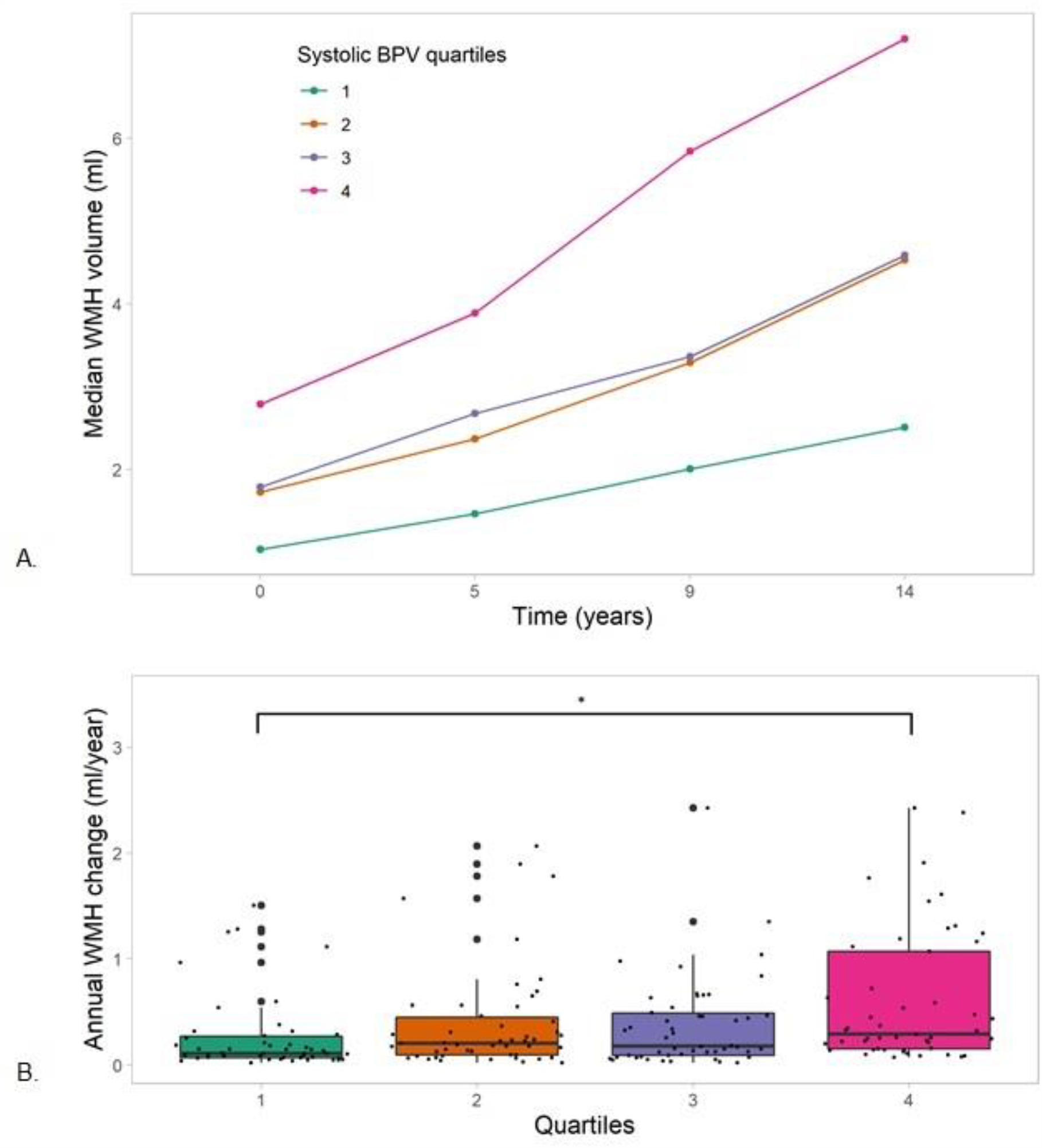
WMH volume changes over 14 years per systolic BPV quartile. A) Median WMH volume (ml) per systolic BPV quartile at each visit. B) Rate of WMH change (ml/year) per systolic BPV quartile, estimated using mixed-effects modelling (Model A). The boxes map to the median, 25^th^ and 75^th^ quartiles. * p < 0.05. BPV = Blood Pressure Variability, WMH = White Matter Hyperintensity.

### BPV and risk of incident lacunes

Incident lacunes were observed in 30 of 199 participants (15.5%). Higher systolic BPV was associated with higher risk of incident lacunes after adjustment for confounders (Incidence Rate Ratio (IRR): 1.09, CI 1.00-1.19, p = 0.040). Risk of incident lacune at any timepoint was significantly higher in systolic BPV Q3 (IRR: 5.76, CI 1.40 – 39.32, p = 0.034) and Q4 (IRR: 5.29, CI 1.26 – 36.59, p = 0.046) compared with Q1 (Table 2). Diastolic BPV was not associated with risk of incident lacunes (IRR: 1.07, CI: 0.97 – 1.18, p = 0.15) (Table S1).

### BPV and MRI markers after 14 years

There was no association between systolic BPV and GM volumes, WM volumes or PSMD at visit 4 (Table 3). Moreover, systolic BPV was not associated with increased risk of microbleeds at visit 4 (Table 3). Diastolic BPV was associated with lower WM volumes in participants in Q2, but no other associations with GM and WM volumes, PSMD and microbleed count were observed (Table S2).

**Table 3.** Association between systolic BPV and MRI markers of SVD at visit 4. Data represent β estimates (95% CI), IRR values (95% CI) or p-values. *Log transformed. GMV: Grey Matter Volume, IRR: Incidence Rate Ratio, PSMD: Peak Skeleton of Mean Diffusivity, WMV: White Matter Volume, WMH: White Matter Hyperintensity.

| Systolic BPV by quartile |  |  |  |  |  |  |  | Continuous systolic BPV |  |
| --- | --- | --- | --- | --- | --- | --- | --- | --- | --- |
|  | Q1 | Q2 |  | Q3 |  | Q4 |  |  |  |
| | | Standardized $\beta$<br>(95% CI) | P<br>value | Standardized $\beta$ (95% CI) | P<br>value | Standardized $\beta$ (95% CI) | P<br>value | Standardized $\beta$ (95% CI) | P<br>value |
| **GMV\*** | Ref | -0.02<br>(-0.43 – 0.30) | 0.90 | 0.01<br>(-0.32 – 0.34) | 0.96 | -0.12<br>(-0.46 – 0.22) | 0.50 | -0.04<br>(-0.16-0.07) | 0.47 |
| **WMV\*** | Ref | 0.28<br>(-0.11-0.67) | 0.16 | 0.31<br>(-0.08 – 0.7) | 0.12 | 0.25<br>(-0.15 – 0.66) | 0.22 | 0.03<br>(-0.12-0.17) | 0.70 |
| **PSMD\*** | Ref | 0.09<br>(-0.25 – 0.44) | 0.60 | 0.16<br>(-0.19 – 0.51) | 0.38 | 0.34<br>(-0.02 – 0.70) | 0.07 | 0.08<br>(-0.05 – 0.20) | 0.24 |
| **Microbleed count** |  | IRR<br>(95% CI) | P<br>value | IRR<br>(95% CI) | P<br>value | IRR<br>(95% CI) | P<br>value | IRR<br>(95% CI) | P<br>value |
|  | 1.00 | 0.74<br>(0.32-1.68) | 0.47 | 0.83<br>(0.37-1.86) | 0.66 | 2.01<br>(0.87-4.65) | 0.09 | 1.03<br>(0.97 – 1.10) | 0.33 |

## Discussion

We found that higher systolic visit-to-visit BPV was associated with increased progression of WMH volume and higher risk of incident lacunes over 14 years in patients with SVD.

The cross-sectional association between long-term BPV and SVD markers has recently been evaluated in two meta-analyses. Both studies concluded that higher systolic BPV is associated with higher odds for SVD, while data for diastolic BPV were lacking. Evidence on the longitudinal association between BPV and SVD progression is scarce. Our findings further support the link between systolic BPV and progression of WMH but over a period of 14 years.^14,15^ This association was especially present in the participants in the highest BPV quartile and remained significant after adjusting for baseline WMH volume, one of the strongest predictors for WMH progression.^22^ Moreover, we found an increased risk of incident lacunes over 14 years in patients with higher BPV, which has not been described previously. This may be explained by variability in method used to assess incident lacunes. Previous investigators compared measurements from independent readers instead of side-by-side rating of repeated MRI scans.^15^ In our study, we made use of difference images, which reduced the risk of measurement errors. This method therefore offered more reliable assessment of incident lacunes than manual rating. There was no clear association between diastolic BPV and SVD burden or progression in this study. Most studies examining cerebrovascular damage and BPV mainly report associations with systolic BPV and the prognostic value of diastolic BPV remains to be determined.^12,23^ The different outcomes reported for systolic and diastolic BPV and suggest that different pathophysiological mechanisms may be at play. It can be hypothesized that systolic BPV is a reflection of aging and vascular stiffness, while diastolic BPV reflects autonomic dysfunction.^24^

The effects of BPV on white matter microstructural integrity are less commonly investigated and the results are inconsistent across studies.^25^ We calculated PSMD, a DTI marker that is suggested to be more sensitive to SVD-related brain injury than other common SVD MRI markers and other DTI parameters (i.e. Fractional Anisotropy (FA) and Mean Diffusivity (MD)).^21,26^ The lack of an association between BPV and PSMD in this study may be due to attrition bias, because the participants with the highest BPV likely had a higher SVD burden and may not have completed all follow-ups. Larger studies are needed to assess whether increased BPV is associated with lower microstructural integrity.

The mechanisms that link BPV and small vessel damage remain poorly understood. Animal studies suggest that large BPV may cause endothelial dysfunction by inhibiting nitric oxide production, which leads to an imbalance in vascular homeostasis.^27 28^ Furthermore, larger variations in BP cause mechanical stress on vessel walls, which triggers several structural changes that may contribute to greater arterial stiffness. ^21^ Increased arterial stiffness is suggested to cause increased pulsation of blood flow and less dampening of blood flow, and these processes may cause microstructural damage.^29^ The involvement of arterial stiffening is confirmed by a recent post-mortem study, where greater BPV was associated with a higher burden of atherosclerosis of the circle of Willis and arteriolosclerosis.^30^ It is important to note that the link between BPV and SVD could also reflect reverse causality, as pre-existing vascular damage may disturb central autonomic autoregulation, resulting in larger BPV.^31^ In this study, systolic BPV was already associated with WMH volumes at baseline, supporting the hypothesis of reverse causality. However, this must be interpreted with caution, since we cannot rule out that BPV was already elevated prior to study participation. This finding emphasizes the importance of correcting for pre-existing SVD when examining the association between BPV and SVD.

It could be argued that BPV is higher in hypertensive patients and the observed association between BPV and cerebral damage is therefore mediated by mean BP levels. We adjusted all our analyses for mean BP to reduce the effect of BP levels. Our findings further support the recent meta-analysis in which it was also concluded that BPV is associated with SVD burden independent of mean BP.^23^ Also in the SPRINT-MIND trial, higher BPV was associated with higher risk of dementia despite excellent BP control.^32^ This growing body of evidence highlights the clinical significance of BPV.

This is the first study that examines the effects of BPV on SVD progression over a 14-year time-period in a large cohort of participants with SVD. Because we only included participants with four extensive imaging assessments, we were able to accurately model progression of WMH over time. We used difference images to reliably assess incident lacunes and semi-automatic quantification to examine WMH volumes, which is more reliable to assess progression over different scans. BP was measured three times at each visit, which reduces risk of measurement error. Nevertheless, some limitations need to be addressed. First of all, it is important to note that the interval between BP measurements used in this study is relatively long (3.4-5.4 years). Although this is common in BPV studies, long-term BPV might be affected by other factors, such as changes in antihypertensive treatment over time, seasonal BP changes, medication adherence and inaccuracy of BP measurements.^33^ However, visit-to-visit BPV over years has still been shown to have prognostic value.^34^

Another limitation are the changes in scanner and protocol during the follow-ups, possibly leading to measurement error of (progression of) WMH volume. This is almost inevitable in long follow-up studies due to technical advances over the years and we accounted for this by including MRI system as a term in our models. Furthermore, nonparticipants were older, had more vascular risk factors and had a higher SVD burden at baseline in this study. Since age and WMH burden are strong predictors for WMH progression, loss-to-follow-up of these participants could have led to an underestimation of the true WMH progression.^22^

If a causal relationship between BPV and SVD is established, lowering BPV may be a target to prevent or mitigate SVD. It has been suggested that calcium channel blockers and non-loop diuretics reduce visit-to-visit BPV in addition to lowering mean BP levels.^35^ Moreover, a healthy lifestyle was associated with lower BPV in young and healthy adults, emphasizing the importance of lifestyle modification in patients at risk of cerebrovascular disease.^36^ However, this evidence is limited and randomized controlled trials are needed to examine if pharmacological or lifestyle modifications can be used to target elevated BPV and related vascular damage.

In conclusion, this longitudinal observational study suggests that higher BPV is associated with more WMH progression and more incident lacunes over 14 years in SVD patients. If this association is causal, reducing BPV could be a target to slow down SVD progression and reduce risk of dementia and stroke. Future studies are needed to confirm our findings and examine the temporal order of the observed association.

## Data Availability

Data is available upon reasonable request.

## Non-standard Abbreviations and Acronyms

AIC: Akaike Information Criterion
BP: Blood Pressure
BPV: Blood Pressure Variability
CSF: Cerebrospinal Fluid
CV: Coefficient of Variation
DTI: Diffusion Tensor Imaging
DWI: Diffusion Weighted Imaging
FA: Fractional Anisotropy
GM: Grey Matter
FLAIR: Fluid Attenuated Inversion Recovery
ICV: Intracranial Volume
LME: Linear-Mixed Effects
MD: Mean Diffusivity
MP2RAGE: Magnetisation Prepared 2 Rapid Acquisition Gradient Echoes
MRI: Magnetic Resonance Imaging
PSMD: Peak Width of Skeletonized Mean Diffusivity
RUNDMC: Radboud University Nijmegen Diffusion tensor Magnetic resonance imaging Cohort
STRIVE: Standards for ReportIng Vascular Changes on nEuroimaging
SVD: Small Vessel Disease
SWI: Susceptibility-weighted image
WM: White Matter
WMH: White Matter Hyperintensities

## Acknowledgments

None.

